# Associations of Very Low Lipoprotein(a) Levels With Risks Of New-Onset Diabetes And Non-Alcoholic Liver Disease

**DOI:** 10.1101/2023.07.10.23292471

**Authors:** Ming Wai Yeung, M. Abdullah Said, Yordi J. van de Vegte, Niek Verweij, Robin P.F. Dullaart, Pim van der Harst

**Author notes:** Address for correspondence: Ming Wai Yeung and Pim van der Harst, Department of Cardiology, University of Groningen, University Medical Center Groningen, Hanzeplein 1, 9700 RB Groningen, The Netherlands, and. Contributed equally (Robin P.F. Dullaart, Pim van der Harst).

## Abstract

**Background and aims:** We aimed to study the association of very low serum Lipoprotein(a) [Lp(a)] concentrations with new-onset type 2 diabetes (T2D) and non-alcoholic liver disease (NAFLD) in the context of statin usage in the UK Biobank, a large prospective population cohort.

**Methods:** Using an extended biomarker dataset, we identified 47,362 participants with very low Lp(a) concentrations (<3.8 nmol/L) from a total of 451,479 participants. With a median follow-up of 12.3 years, we assessed the risk of new-onset cardiometabolic diseases in participants stratified by statin usage with Cox proportional hazards models. We performed two-sample Mendelian randomization MR analyses to test causal relationship between genetically predicted Lp(a) and T2D and NAFLD.

**Results:** Taking the participants with Lp(a) within reportable range as the reference group, the hazard ratios (HR) for T2D were 1.06 (95% confidence interval, CI 1.01-1.12) and for NAFLD 1.35 (95% CI 1.26-1.45) respectively for participants with very low Lp(a) (<3.8 nmol/L). The risk for new-onset T2D was higher in participants using statin (adjusted HR 1.14; 95% CI 1.05-1.24). This was contrasted by higher risk of NAFLD in participants not on statin (HR 1.40; 95%CI 1.28-1.53). There was no evidence for causal links between genetically predicted Lp(a) and T2D nor NAFLD in two-sample MR analyses.

**Conclusions:** Very low Lp(a) was associated with higher risks of T2D and NAFLD in a prospective analysis of the UK Biobank. These associations were influenced by lipid lowering medication usage. MR analyses did not support causality for these inverse associations.

## Introduction

Lipoprotein(a) [Lp(a)] is a specific lipoprotein produced in the liver that consists of a polymorphic glycoprotein, called apolipoprotein(a) [apo(a)] linked to apolipoprotein-B(100) on an LDL particle in a 1:1 ratio. Ample evidence supports a causal association between elevated Lp(a) and various cardiovascular diseases (CVD) [1–3]. Interestingly, recent epidemiological studies raised the possibility that very low Lp(a) is associated with increased risk of new-onset type 2 diabetes mellitus (T2D). [1]. However, it is unclear whether Lp(a) is a mere marker or whether there is a causal mechanism behind this possible inverse relationship. An inverse association has also been suggested for fatty liver disease [4]. Closely linked to metabolic dysfunction, the causal relationship between non-alcoholic liver disease (NAFLD) (new nomenclature: metabolic associated steatotic fatty liver disease [MASLD]) and diabetes has been supported by evidence from both epidemiological studies and animal studies [5–7]. The opposed direction of the Lp(a) association with new-onset T2D, NAFLD and CVDs also raises the question whether the risk of diabetes might be modified by statin treatment [8].

In the current study, we aimed to investigate Lp(a) associations with new-onset T2D and NAFLD in the UK Biobank, a large population-based cohort, and examined effect modification by treatment with statin. We further examined the potential causal relationship of Lp(a) with T2D and NAFLD using a two-sample MR approach.

## Methods

### Study Population

The UK Biobank study is a population-based prospective cohort in the United Kingdom in which over 500,000 participants aged between 40 and 69 years were included between 2006-2010. All participants have given informed consent for this study. The UK Biobank has ethical approval from North West - Haydock Research Ethics Committee (REC reference: 16/NW/0274). Details of the UK Biobank study has been described in detail previously [9]. This research has been conducted using the UK Biobank Resource under Application Number 74395. Demographics including age, sex, smoking status and ethnic background were collected at the baseline visit.

### Ascertainment of health outcomes

Ascertainment of diseases and medication usage in the UK Biobank was based on a combination of hospital inpatient records, primary care records and self-reported records during an interview with a trained staff (**Table S1**). Briefly, T2D diagnosis was captured using ICD10 code E10-E14 or equivalent diagnosis codes; NAFLD diagnosis was captured using K721, K740, K741, K742, K746, K758 and K760 or equivalent diagnosis codes [10]. Participants with prevalent disease at baseline per outcome were excluded from regression analysis. For T2D, we additionally excluded participants with baseline serum glycated haemoglobin (HbA1c) ≥6.5% (48 mmol/mol). Health outcome data were processed and extracted using the ukbpheno v1.0 package in R [11]. Participant follow-up started at inclusion and ended at date of event, death or last recorded follow-up (**Table S2**), whichever occurred first.

### Serum biomarker measurement and extreme Lp(a) concentration

Lp(a) (in nmol/L) was measured using an immuno-turbidimetric assay (Randox Bioscience, UK). The reportable range of the assay is 3.8-189 nmol/L. To identify participants with extreme Lp(a) concentration, we obtained Lp(a) concentrations outside of the reportable range from the UK Biobank (Return 2321). We therefore considered participants to have very low Lp(a) if they had Lp(a) concentrations <3.8 nmol/L; and those with Lp(a) concentrations >189 nmol/L very high Lp(a) (**Figure 1**).

**Figure 1.**
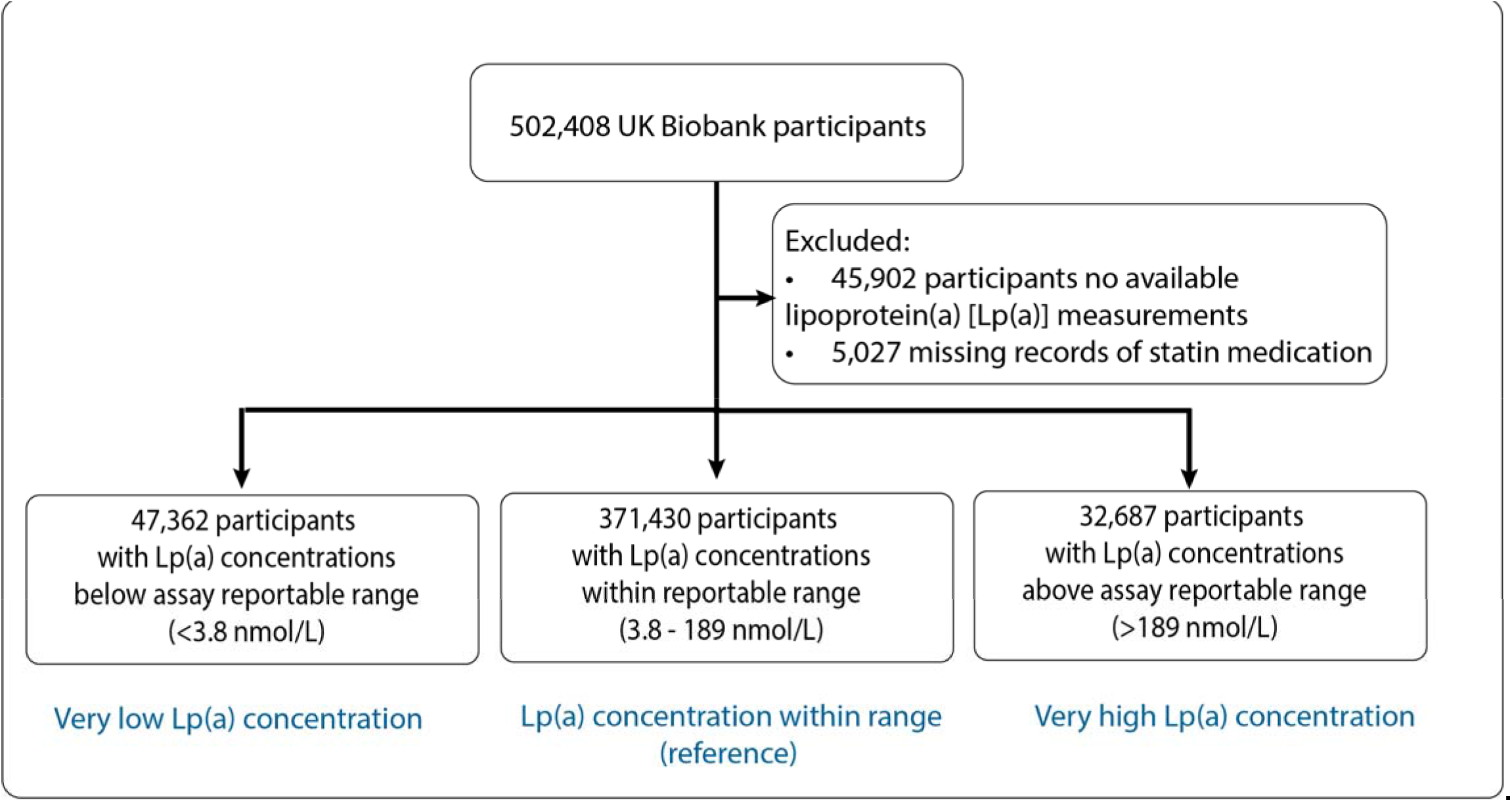
Study population.

Non-fasting venous blood samples were drawn during participants’ baseline visit to the Assessment Centre [12]. Low density lipoprotein cholesterol (LDL-C) was measured using an enzymatic selective protection method while high density lipoprotein cholesterol (HDL-C) was measured using an enzyme immuno-inhibition method. Triglycerides were measured using an enzymatic method, namely the glycerol-3-phosphate (GPO)-peroxidase (POD) chromogenic method. All biomarkers were measured on a Beckman Coulter AU5800 (Beckman Coulter [UK], Ltd). Glycated haemoglobin (HbA1c) was measured in mmol/mol with high performance liquid chromatography method (Bio-Rad Laboratories, Inc.) on Bio-Rad Variant II Turbo analysers[13].

### Statistical analyses

Data are presented as median (IQR) for continuous variables and as counts with percentages for discrete and categorical variables, respectively.

#### Cox regression analyses

We performed two Cox proportional hazards regression analyses to test the association of Lp(a) with coronary artery disease (CAD), as compared to T2D and NAFLD separately, stratified by statin use. We first examined the disease risk by Lp(a) categories. We estimated HRs with 95% confidence intervals in participants with either very low or very high Lp(a) concentrations against participants with Lp(a) concentrations between 3.8 and 189 nmol/L. In the second analysis, we directly modelled the inverse rank normalized serum Lp(a) concentrations linearly only in participants with Lp(a) in the reported range. In both analyses, we considered four models. Model 1 was adjusted for age at baseline and sex; model 2 was further adjusted for body mass index (BMI) and hypertension; model 3 was further adjusted for LDL-C and HDL-C; and finally model 4 was further adjusted for triglycerides. We additionally examined if these associations were affected by use of statin, a lipid-lowering medication with reported link to increased risk of diabetes[14]. Cox regressions were performed using Stata 17.1 (StataCorp LLC). We considered results with *p*<0.001 to be statistically significant to correct for multiple testing.

#### MR analyses

We performed two-sample Mendelian randomization (MR) analyses to assess potential causal effects of Lp(a) on T2D and NAFLD. We took the lead SNPs identified in the UK Biobank GWAS as instrumental variables for Lp(a) [3]. We took estimates from the summary statistics data of the DIAMANTE Consortium (1,159,055 controls and 180,834 [13.5%] cases) for diabetes [15] and EPoS Consortium for NAFLD (17,781 controls and 1,483 [7.7%] histologically confirmed cases)[16]. SNPs (genetic instruments) that were not available in the outcome GWAS were replaced with proxies in linkage disequilibrium (LD) of r2>0.8 respectively or excluded from the MR if no eligible proxies were available. Harmonization of SNP effects and the MR analyses were performed using R (version 4.0.3), the TwoSampleMR package (version 0.5.6) [17], MendelianRandomization (version 0.6.0)[18], MRMix (version 0.1.0)[19], MR-PRESSO (Pleiotropy Residual Sum and Outlier) (version 1.0)[20] and mr.raps (version 0.2)[21].

MR estimates were obtained using an inverse variance weighted (IVW)-fixed effects model followed by sensitivity analyses to assess the robustness of the findings of the IVW estimates. For this, we used IVW-random effects, MR-Egger[22], median-, mode-based estimator MR analyses, MR-PRESSO, MR-Lasso[23], MR-Mix and MR-RAPS. Weak instrument bias was tested using F-statistics[24], with F-statistics >10 indicating low risk of weak instrument bias. SNPs with F-statistics ≤10 were excluded from the analyses. The I2 index (> 25%)[25] and Cochran’s Q statistic (P < 0.05) [26] were determined to assess potential heterogeneity within IVW-fixed effects estimates, indicating at least balanced horizontal pleiotropy for some of the SNPs.

The MR-Egger method allows for a non-zero intercept hence capturing the unbalanced horizontal pleiotropic effects, if these pleiotropic effects are independent of their association with the exposure of interest (Instrument Strength Independent of Direct Effect [InSIDE] assumption)[27]. Rücker’s Q’was calculated to assess heterogeneity within MR-Egger analyses, and the difference between Rücker’s Q’ statistic and Cochran’s Q statistic (Q-Q’) was tested [26]. The MR-Egger intercept was tested for a deviation from zero. We consider presence of unbalanced horizontal pleiotropy in the in the case of a significant Q-Q’ and a non-zero MR-Egger intercept. The MR-Egger estimate was considered the causal estimate rather than the MR-IVW method in presence of unbalanced horizontal pleiotropy if the general InSIDE assumption holds (**Figure S1**).

We additionally assessed the potential weak instrument bias in the MR-Egger analyses by calculating the I^2^_GX_, which is the true variance of the SNP-exposure association and indicates low risk of measurement error if >95%[27].

The median/mode-based methods take, respectively, the median or mode of the ratio estimates instead of the weighted mean in IVW method. They are therefore robust against the presence of a limited number of invalid instruments. MR-PRESSO method performs the IVW method after exclusion of SNPs with MR estimates that differ substantially from the rest, thereby adjusting for the potential pleiotropy[20]. Similarly, the MR-Lasso method is another augmented version of the IVW method with outliers removed. MR-RAPS and MR-Mix methods adjust the estimates to address the violation of valid instrument assumption.

## Results

We studied 451,479 participants (54.3% women with mean age 57.1 years) with Lp(a) measurements and information on statin usage. Among these participants, 371,430 had Lp(a) measurements within the reportable range of 3.8 −189 nmol/L (median 21.1; IQR 9.6-61.9); 47,362 participants had very low Lp(a) concentrations (<3.8 nmol/L) while 32,687 participants had very high Lp(a) (>189 nmol/L). A total of 83,405 (18.5%) participants were on statin at baseline. Participants with very low Lp(a) concentrations were more likely to be men and to be White, had lower LDL-C and HDL-C but higher triglycerides (**Table 1**).

**Table 1.** Baseline characteristics of UKB participants by Lp(a) concentration.

| Variable | Lp(a) <3.8 nmol/L | 3.8≤ Lp(a) ≤189 nmol/L | Lp(a) >189 nmol/L | p |
| --- | --- | --- | --- | --- |
| N | 47,362 | 371,430 | 32,687 |  |
| Age, years (IQR) | 57.57 (49.69, 63.43) | 58.24 (50.48, 63.71) | 59.71 (52.77, 64.31) | <0.001 |
| Sex |  |  |  | <0.001 |
| Men | 24734 (52.2%) | 168787 (45.4%) | 12612 (38.6%) |  |
| Women | 22628 (47.8%) | 202643 (54.6%) | 20075 (61.4%) |  |
| Ethnicity |  |  |  | <0.001 |
| White | 46110 (97.4%) | 348537 (93.8%) | 30811 (94.3%) |  |
| Asian | 545 (1.2%) | 9224 (2.5%) | 447 (1.4%) |  |
| Black | 86 (0.2%) | 6094 (1.6%) | 879 (2.7%) |  |
| Mixed | 167 (0.4%) | 2315 (0.6%) | 197 (0.6%) |  |
| Other/Unknown | 454 (1.0%) | 5260 (1.4%) | 353 (1.1%) |  |
| Current smoker | 5054 (10.7%) | 39389 (10.6%) | 3389 (10.4%) | 0.35 |
| BMI, kg/m<sup>2</sup> | 27.09 (24.30, 30.48) | 26.69 (24.10, 29.81) | 26.89 (24.38, 30.00) | <0.001 |
| Systolic blood pressure, mmHg | 133.03 (121.55, 145.47) | 131.71 (120.21, 144.47) | 133.05 (121.13, 145.50) | <0.001 |
| Diastolic blood pressure, mmHg | 82.13 (76.47, 88.19) | 81.72 (76.06, 87.39) | 81.73 (76.06, 87.79) | <0.001 |
| LDL-C, mg/dl | 128.83 (107.49, 151.45) | 135.75 (113.91, 158.68) | 145.30 (120.37, 170.71) | <0.001 |
| HDL-C, mg/dl | 52.74 (43.85, 63.64) | 54.02 (45.28, 64.65) | 56.30 (47.37, 66.93) | <0.001 |
| Triglycerides, mg/dl | 136.67 (94.33, 201.33) | 130.82 (92.29, 189.37) | 127.19 (91.59, 180.87) | <0.001 |
| Hypertension | 15112 (32.0%) | 113374 (30.6%) | 12168 (37.3%) | <0.001 |
| Statin usage | 8477 (17.9%) | 65177 (17.5%) | 9751 (29.8%) | <0.001 |
| New-onset T2D | 5189 (11.0%) | 32079 (8.7%) | 3076 (9.4%) | <0.001 |
| New-onset non alcoholic fatty liver disease | 1102 (2.3%) | 5586 (1.5%) | 429 (1.3%) | <0.001 |
| New-onset coronary artery disease | 4449 (9.4%) | 36438 (9.8%) | 5005 (15.3%) | <0.001 |
| New-onset aortic stenosis | 424 (0.9%) | 3662 (1.0%) | 641 (2.0%) | <0.001 |
| New-onset heart failure | 1603 (3.4%) | 11984 (3.2%) | 1404 (4.3%) | <0.001 |
| New-onset stroke | 1993 (4.2%) | 15627 (4.2%) | 1719 (5.3%) | <0.001 |

BMI: body mass index, HDL-C: high density lipoprotein cholesterol, IQR: interquartile range, LDL-C: low-density lipoprotein cholesterol.

During a median follow-up of 12.3 years (IQR 11.5-13.1) 40,344 participants were diagnosed with T2D and 7,117 with NAFLD. There were 45,892 cases of new-onset CAD. There was more new-onset diabetes (11.0% vs. 8.7% vs. 9.4%) as well as NAFLD (2.3% vs. 1.5% vs. 1.3%) among participants with very low Lp(a) concentrations (**Table 1**).

In all participants combined, Cox regression analyses showed that very low Lp(a) concentrations at baseline showed trend of increased risk of T2D but the association was not statistically significant after considering multiple testing corrections (fully adjusted HR:1.06; 95% CI 1.01-1.12; *p*=0.016). We observed increased risk of NAFLD (fully adjusted HR:1.35; 95% CI 1.26-1.45; *p*<0.001), in contrast to the decreased risk of CAD (fully adjusted HR 0.86; 95% CI 0.83-0.90; *p*<0.001) (**Figure 2**). This inverse association with new-onset T2D was more prominent in participants using statin at baseline (age- and sex-adjusted interaction *p*=0.04). Additional adjustments attenuated the association and it lost significance after further adjustment for triglycerides, both among participants on statin (HR 1.14; 95% CI 1.05-1.24; *p*=0.003) and in those not on statin (HR 1.06; 95% CI 1.00-1.13; *p*=0.055) respectively (**Table S3**). The association with NAFLD remained statistically significant in the fully adjusted model in whole cohort (**Figure 2**). Subgroup analysis revealed a slightly higher point estimate of the HR in participants not using statin (**Table S4**). Reverse trends were observed in participants with very high Lp(a) concentrations (**Figure S2**).

**Figure 2.**
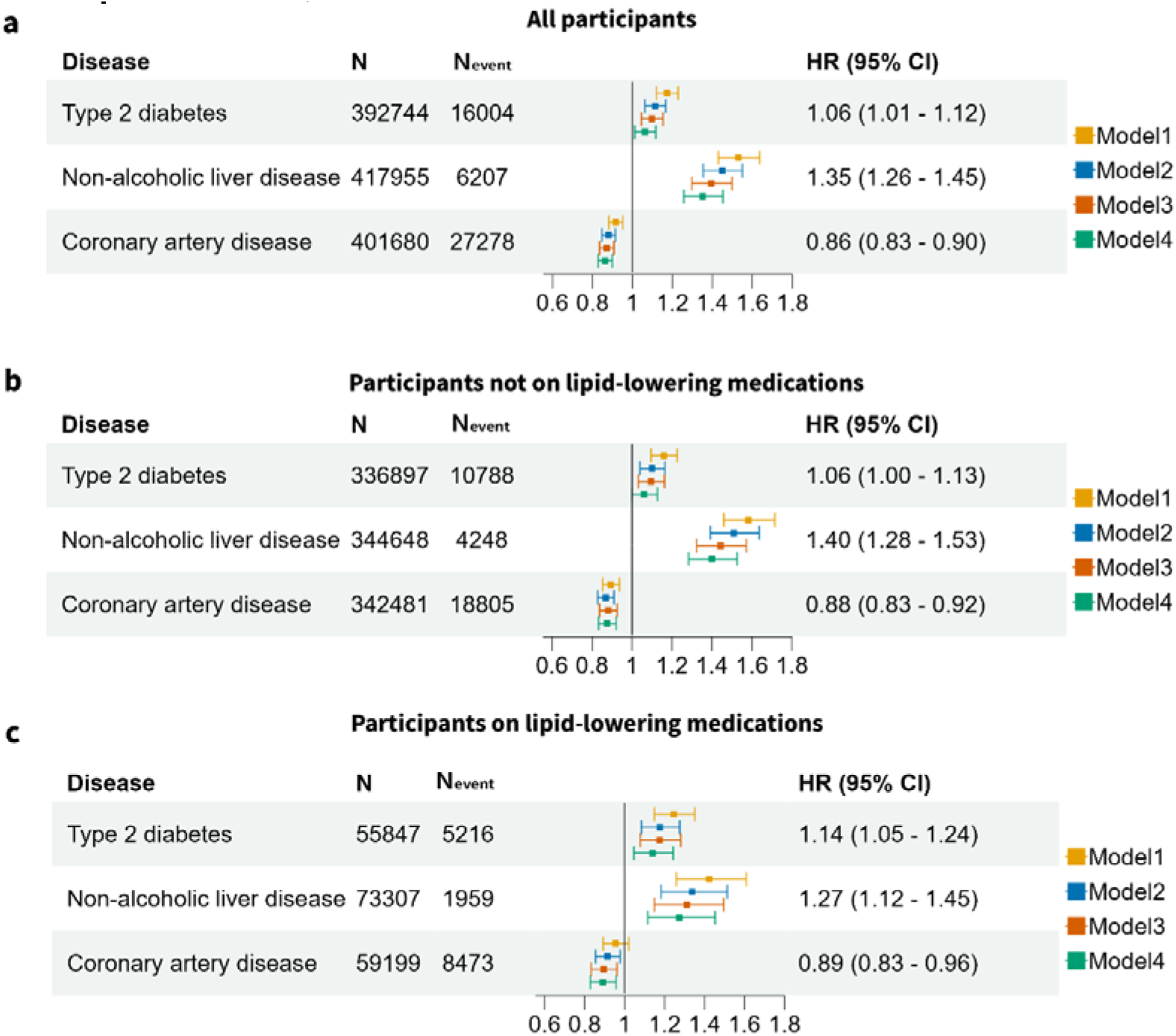
Forest plot illustrating the unadjusted and adjusted hazard ratios of very low Lp(a) concentrations at baseline with the development of T2D, NAFLD and CAD. Hazard ratios for diseases in **(a)** all participants **(b)** participants not using statin and **(c)** participants using statin in model 4. Model 1: adjusted for age and sex; Model 2: Model 1 + body mass index and hypertension; Model 3: Model 2 + low-density lipoprotein cholesterol and high-density lipoprotein cholesterol; Model 4: Model 3 + triglycerides. Hazard ratios (HR) with 95% confidence intervals (CI) of Model 4 are shown for new-onset T2D, NAFLD and CAD. N: Number of participants, N_event:_ number of events.

We continued by modeling inverse rank normalized Lp(a) concentrations among participants whose Lp(a) concentrations were within the reportable range (N=371,430). We observed smaller point estimates with neither association with diabetes nor NAFLD being significant in the fully adjusted Model 4 (**Figure S3**). Given the ethnic difference in Lp(a), we assessed the associations with new-onset T2D (**Table S5**) and NAFLD (**Table S6**) by ethnic groups in participants not on statin. We found no evidence of interaction effects between either disease and White ethnicity (age and sex adjusted interaction *p*=0.149 for T2D and *p*=0.456 for NAFLD, respectively).

We next performed two-sample MR analyses to test whether genetically predicted Lp(a) is associated with T2D and NAFLD with GWAS summary statistics from external cohorts namely DIAMANTE for T2D and EPos for NAFLD respectively. A total of 37 and 35 genetic instruments for Lp(a) were available for the MR analyses in the DIAMANTE and EPoS GWAS repositories. We did not find evidence for an association of genetically predicted Lp(a) with T2D (IVW-fixed effects OR:1.02; 95% CI: 0.99-1.04, *p*=0.149) (**Figure 3**). Sensitivity analyses indicated presence of balanced vertical pleiotropy with non-significant intercept (−0.0049 ± 0.0030, *p* = 0.116) in the MR-Egger model. Estimates by IVW-random effects (OR:1.02; 95% CI: 0.96-1.08, *p*=0.564) were consistent with most methods except for the MR-Mix model. We furthermore found no evidence for an association of genetically predicted Lp(a) with NAFLD (IVW-fixed effects OR:1.00; 95% CI: 0.96-1.03, *p*=0.825) with evidence for horizontal pleiotropy in the sensitivity analyses (**Figure 4**). Estimates in the sensitivity analyses were consistent with IVW-fixed effects models.

**Figure 3.**
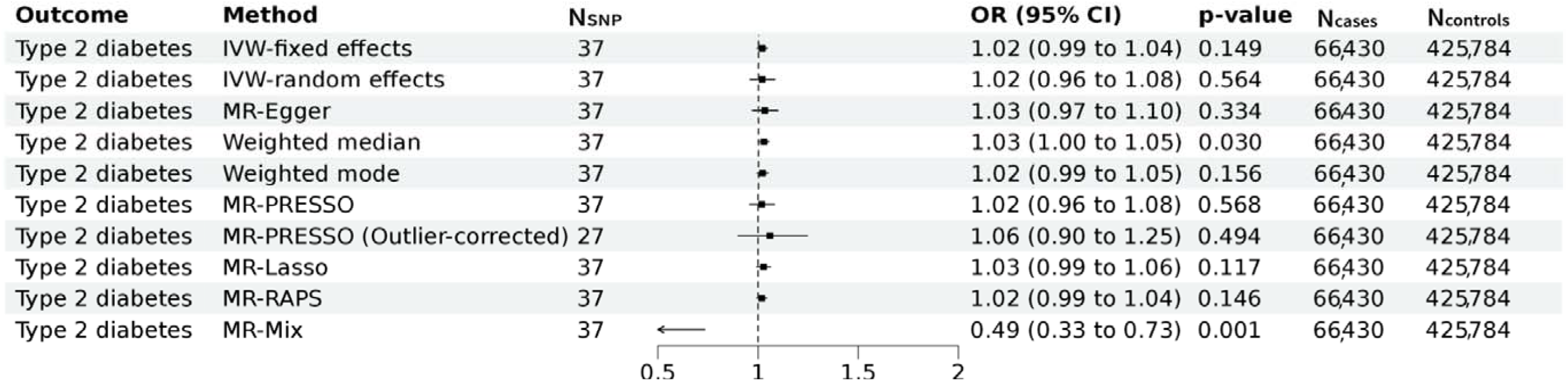
Mendelian randomization assessing the genetic association of Lp(a) with Type 2 diabetes. Forest plot of the Mendelian randomization results of Lp(a) with type 2 diabetes. Results of the MR IVW-fixed effects, IVW-random effects, MR Egger, weighted median, weighted mode, MR-PRESSO, MR Lasso, MR-Raps and MR-mix are provided. Odds ratios and 95% confidence intervals are shown. We found evidence for horizontal pleiotropy in the current analyses (I^2^ index 84.1% (CI= 78.9-88.0); Cochran’s Q = 230, degrees of freedom (df) = 36, *p* =2.4×10^−29^; Q-Q’ = 16, df = 1, *p* = 8×10^−5^; MR-Egger intercept −0.0049 ± 0.0030, *p* = 0.116). There was no evidence for weak instrument bias in the MR-Egger regression (I^2^_GX_ = 0.99).MR: Mendelian randomization, IVW = inverse variance weighted, MR-PRESSO: Mendelian Randomization Pleiotropy RESidual Sum and Outlier, MR-RAPS: Mendelian randomization using the robust adjusted profile score, OR = odds ratio, CI = Confidence interval.

**Figure 4.**
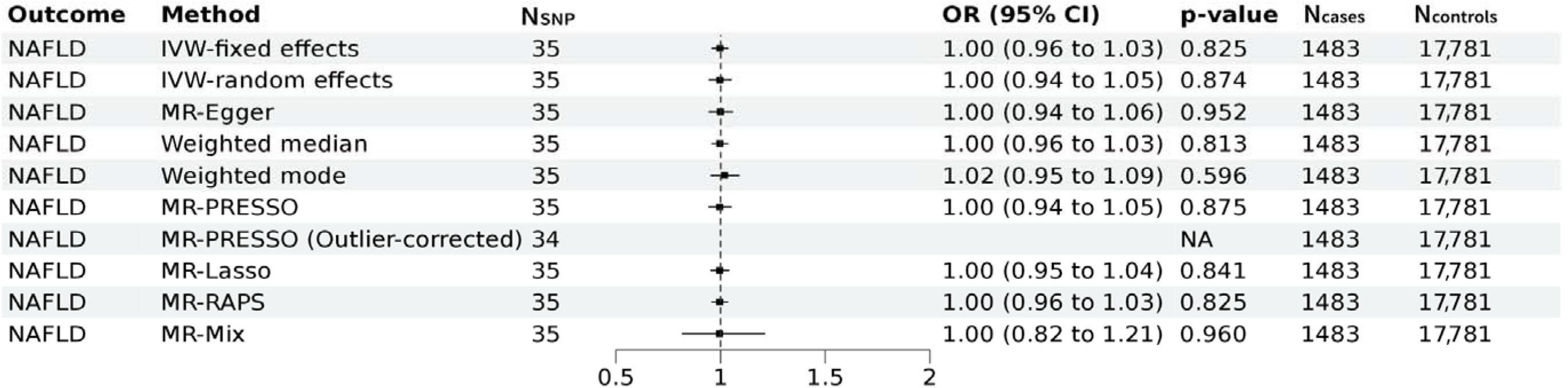
Mendelian randomization assessing the genetic association of Lp(a) with NAFLD. Forest plot of the Mendelian randomization results of Lp(a) with non-alcoholic fatty liver disease (NAFLD). Results of the MR IVW-fixed effects, IVW-random effects, MR Egger, weighted median, weighted mode, MR-PRESSO, MR Lasso, MR-Raps and MR-mix are provided. Odds ratios and 95% confidence intervals are shown. We found evidence for horizontal pleiotropy in the current analyses (I^2^ index 48.6% (CI= 23.8-65.3); Cochran’s Q = 66, degrees of freedom (df) = 34, *p* =0.0008; Q-Q’ = 0.19, df = 1, *p* =0.661; MR-Egger intercept −0.0021 ± 0.0069, *p* = 0.759). There was no evidence for weak instrument bias in the MR-Egger regression (I^2^_GX_ = 0.99).MR: Mendelian randomization, IVW = inverse variance weighted, MR-PRESSO: Mendelian Randomization Pleiotropy Residual Sum and Outlier, MR-RAPS: Mendelian randomization using the robust adjusted profile score, OR = odds ratio, CI = Confidence interval.

## Discussion

Previous work equivocally suggested that higher Lp(a) concentration may be associated with a lower risk of diabetes [1, 28]. Accordingly, the EAS consensus statement published in 2022 has suggested that there is a unmet need to dissipate whether a potential association of very low levels of Lp(a) with higher risk of diabetes development is causal[1]. By using an extended biomarker dataset in over 450,000 UK Biobank participants, we confirmed epidemiological observation of the higher risk of T2D in individuals with very low Lp(a) concentrations (<3.8 nmol/L) contrasting its protective effect on CVDs. We however found that these associations were no longer significant after adjusting for other cardiometabolic risk factors. In contrast, association between low Lp(a) and increased NAFLD risk remained in the fully adjusted model.

Given previous report of increased risk of new-onset T2D with statin use[14], we further assessed the association in subgroups stratified by statin. Notably, we found that the risk of diabetes was enhanced in individuals on statin while the risk of NAFLD was attenuated in the context of statin. However the statistical interactions were not significant as assessed by a product term in the Cox model.

Our results from the two sample MR analyses using 37 lead SNPs, which included rs10455872 (a common intronic SNP strongly associated with Lp(a) concentration in White individuals), as genetic instruments, did not support a causal relationship between Lp(a) and T2D nor NAFLD. Our results extend findings of previous work which solely used rs10455872 [29]. Sensitivity analyses suggested presence of pleiotropy in the association between Lp(a) with both traits though we found the estimates remained consistent in the sensitivity analyses. Association between Lp(a) isoform size and risk of T2D were also reported [30] though we were unable to investigate the role of Lp(a) isoform size as these data are not available in the UK Biobank, which is among the limitations of the current study.

Several studies have implicated a role of triglyceride-rich lipoproteins in the metabolism of Lp(a) [8, 31, 32], whereas conversely genetically predicted Lp(a) may influence very low density lipoprotein (VLDL) metabolism [33]. These findings corroborate to study the association of Lp(a) with new-onset T2D and NAFLD in the context of statin use, and to adjust for triglycerides when examining the association of Lp(a) with T2D and NAFLD. Experimental studies are needed to study the intricate interactions between Lp(a) and VLDL as well as its involvement in the associations with diabetes and non-alcoholic liver disease reported in observational studies.

Strengths of the current study Include the large dataset from the UK Biobank obviating the need to harmonize plasma Lp(a) measurements. However, most UK Biobank participants are White, precluding to test robust interactions of Lp(a) with ethnicity on diabetes development. This is relevant because the distribution of plasma Lp(a) varies considerably among ethnicities with a median concentration being much higher in Black vs, White individuals[1].

## Conclusions

Using prospective data of the UK Biobank repository, very low association for Lp(a) concentrations with T2D and NAFLD were modified by statin usage. We found no evidence for an association between genetically predicted Lp(a) and T2D or NAFLD. Our current findings in both longitudinal analysis and MR analysis do not substantiate any concerns in exacerbated T2D risk by aggressive Lp(a) lowering therapy.

## Data Availability

Data are available on application to the UK Biobank https://www.ukbiobank.ac.uk/enable-your-research/register

## Declaration

### Ethics approval and consent to participate

The UK Biobank has ethical approval from North West - Haydock Research Ethics Committee (REC reference: 16/NW/0274). All participants have given informed consent.

### Availability of data and materials

The data that support the findings of this study are available from the corresponding authors upon reasonable request. Source data may be requested from the UK biobank.

### CRediT authorship contribution statement

**M.W.Y, M.A.S**., **R.P.F.D**., **P.v.d.H:** Conceptualization; **M.W.Y, M.A.S**., **Y.J.v.d.V, N.V**.: Methodology, Software; **M.W.Y, M.A.S**.: Investigation; **R.P.F.D, P.v.d.H:** Supervision; **M.W.Y** :Writing - Original Draft, review & editing; **M.A.S**., **Y.J.v.d.V, N.V**., **R.P.F.D**., **P.v.d.H:** Writing – Critical review & Editing.

## Acknowledgments

This research has been conducted using the UK Biobank resource under application numbers 74395. We thank all UK Biobank participants for their contributions.

