## Supplement for "Associations of Very Low Lipoprotein(a) Levels With Risks Of New-Onset Diabetes And Non-Alcoholic Liver Disease"

Author affiliations:
[1]University of Groningen, University Medical Center Groningen, Department of Cardiology, 9700 RB Groningen, the Netherlands
[2]Department of Cardiology, Division of Heart & Lungs, University Medical Center Utrecht, University of Utrecht, Utrecht, the Netherlands
[3]Department of Endocrinology, University of Groningen, University Medical Center Groningen, Groningen, The Netherlands.

*Contributed equally

**Contents:**

**Tables S1-S2**

**Figures S1-S3 including figure legends**

### Table S1. Clinical codes used to define disease and medication in the UK Biobank

| Outcome | ICD-9 / ICD-10 | OPCS-4 | Self-reported fields | READ2 | CTV3 | BNF | DMD |
|---|---|---|---|---|---|---|---|
| Aortic stenosis | I350, I352 | K621, K622, K623 | 20002(1490) | G5411, G5413, G5414, G5415 | G5411, G5413, G5414, X2011, Xa0Ct, X2015 | | |
| Coronary artery disease | 414, 410, 412; I24, I25, Z955, I21, I22, I23, I252, Z951, Z955 | K40, K41, K42, K43, K44, K45, K46, K49, K50, K75 | 20002(1075), 20004(1070, 1095, 1523),6150(1) | G34y1, G34.., G3..., ZV45L, G34z0, ZV458, 793G., 79280, 79281, 79282, 7928y, 7928z, 79292, 7929y, 7929z, 792.., 7A547, 793Gy, 793Gz, 79283 | G34y1, XE0WG, XE2uV, XaC1g, XaG1Q, XaQiY, ZV458, G34.., X200b, Xa1dP, XaLgU, 79280, 79281, 79282, 7928y, 7928z, 79292, 7929y, 7929z, X00tT, X013N, XE0Em, XaLgZ, XaLga, XaMKE | | |
| Diabetes* | 250; E10-E14 | | 20002(1220,1222,1223), 2443(1),6153(3),6177(3) | C10.. | C10.., X40J4, X40J5, X40J6, X40JJ, X40Jj, X40Jl | | |
| Heart failure | 428; I50, I110, I130, I132, Z941, T862 | K02 | 20002(1076), 20004(1098) | 14S3., G2101, G2111, G21z1, G232., G234., G58.., G5800, G5802, G5803, G5810, G582., SP084, SP111, G581., 1O1.., G583., ZV421 | 14S3., G2101, G2111, G21z1, G232., G234., G58.., G5800, G5802, G5803, G5810, G582., SP084, X202k, X202v, X202w, XE2QG, XaIpn, XaWyi, ZV421, X00y3 | | |
| Hypertension | 402, 403, 404, 405, 401; I11, I12, I13, I15, O10, I10 | | 20002(1065, 1072),6150(4),6177(2),6153(2),20003 | G21.., G220., G221., G23.., G24.., G240., G240z, G241., G241z, G24z., L12.., G22.., G22z., G2z.., G2y.., G21z0, G20.., G20z., x01QX | G21.., G220., G221., G23.., G24.., G240., G240z, G241., G241z, G24z., L12.., XE0Uf, XE0Ug, G2z.., G2y.., Xa0lt, Xa3fQ, Xa0kX, XE0Uc, x01QX | | |
| Non-alcoholic liver disease | K721, K740, K741, K742, K746, K758, K760 | | | J61y4, J61y1, J61y3, J61y7, J62y., J61y6, J61y9, J61y8 | J61y1, J61y7, X307C, X307W, X307b, X307v, XE0b5, XaQIT, J61y8, J61y9, J625. | | |
| Peripheral artery disease | 4439; I739 | L37, L381, L383, L384, L391, L392, L395, L48, L49, L50, L51, L52, L53, L541, L542, L544, L56, L57, L58, L59, L60, L62, L631, L632, L635, L638, L639, L653 | 20002(1087, 1067), 20004(1102, 1103, 1108) | G73., 7A26., 7A270, 7A271, 7A276, 7A277, 7A27C, 7A27E, 7A279, 7A27B, 7A27D, 7A280, 7A281, 7A283, 7A28D, 7A28C, 7A28G, 7A40., 7A41., 7A42., 7A43., 7A440, 7A441, 7A45., 7A46., 7A47., 7A48., 7A4A., 7A4B0, 7A4B1, 7A4B2, 7A4B3, 7A4B9, 7A4By, 7A4Bz, 7A502, 7A2., 7A443, 7A4B., 7A433 | Xa0lV, G73z., XE0VP, XE0F9, 7A270, 7A276, 7A27C, 7A280, 7A283, XaMMk, 7A26., X013S, Xa7pt, XaCLU, X015N, 7A277, 7A281, 7A284, Xa9Iv, X013T, XE0FJ, X016D, X015h, XE0FK, X015H, XE0FN, XM1Ih, XE0FQ, 7A440, 7A441, XaDmi, 7A40., 7A41., 7A42., 7A43., 7A443, X013i, X013k, X013m, X016n, X016o, X70XH, XE0FI, XaDmh, XaG1G, X015g, X015f, XM1M6, XE0FV, X015d, XM1Ik, XM1Im, XE0FW, XM1In, XM1Il, X015X, X015b, X015c, XE0FX, X015Y, X015R, X015a, XE0FY, X015V, X015U, X015T, X015W, XM1Ir, XM1Is, XE0FZ, 7A45., 7A46., 7A47., 7A48., 7A49., 7A4A., X013o, X013p, X013q, X013r, X014V, X014W, 7A502, 7A4Bz, 7A4By, XaMNe, 7A4B2, 7A4B0, XM1Iv, XM1It, XM1Iw, XM1Iq, XM1KK, XaIzW, XaE0L, Xa0Fu, Xa0Fr, XM1J1, XM1Iz, XM1Iy, XM1Ix, XE0Fb, X016p, X0159, X015O, 7A279, 7A27B, 7A27D, 7A28C, 7A28D, Xa7pv, Xa7y1, XaCLY, X015e, XM1Io, X015Z, X015S, XM1Ip, XM1Iu, 7A4B1, 7A4B3, X013u, X014h, X015C, XM1J0, Xa0Fz | | |

| Outcome | ICD-9 / ICD-10 | OPCS-4 | Self-reported fields | READ2 | CTV3 | | |
|---|---|---|---|---|---|---|---|
| Stroke | 430,431,434,435,436,4371, 3361, 36231, 36232, 4329, 43301, 43311, 43321, 43331, 43381, 43391; I60, I61, I63, I64, I65, I629, I678, I690, I693, G45, G951, H341, H342, S066 | A052, A053, A054, L343, L351, L353, | 20002(1081,1082,1086,1491,1583),6150(3) | F4236,G65zz,G64z4,G66..,G64..,F4232,G65..,F4238,G64z.,F4239,G668.,G64z2,G60..,F423.,G679.,G61..,G62..,G655.,G673.,G678.,G667.,G623.,G665.,G61X.,G64z1,G622.,G6400,G650.,G64z3,G613.,G605.,G652.,G662.,G621.,G61z.,G664.,G600.,G6740,G60z.,G640.,G663.,G61X0,G6760,G6732,G6733,G6772,F4231,G641.,G671.,G602.,G67z.,70043,G614.,G604.,F423z,G610.,Gyu6F,G64z0,G61X1,G656.,G660.,F4230,G62z.,G65z.,70041,G6510,G676.,G611.,G6731,G617.,G666.,G612.,G6410,G619.,G6711,G657. | X00D1,XE0VK,XaBEC,XE2aB,G65z.,XE0Wy,F4231,Xa1uW,X00D7,Xa0kZ,Xa00J,G664.,XE0VF,G62..,G65y.,G677.,Xa00I,XaBED,G676.,X00D5,XM0rV,G62z.,G61z.,X00D6,G65z0,G673.,Xa01h,X00DV,Xa00K,X00db,Xa01l,Xa3fV,X00DU,XaEIh,Xa01k,G611.,G663.,X00DR,G621.,G6711,70043,X00Dg,G60z.,G6y..,G675.,G612.,Xa84g,Xa01i,G6760,Xa1hE,G671.,G67..,G614.,Xa01j,XaBM4,70041,X00DN,G613.,X00DO,XaBM5,Gyu60,G610.,G601.,Gyu6F,G683.,G6772,Xa0N7,F1611,G6771,Gyu6E | | |
| Use of statin | | | 20003(1140861958,1140881748,1140910652,1140910654,1141188146,1141195196,1141200040,1140861970,1140864592,1141146138,1141146234,1141192410,1141192414,1140888594,1140888648,1140910632) | bxd,bxe,bxg,bxi,bxj,bxk,x01R2,x01R3 | bxd,bxe,bxg,bxi,bxj,bxk,x01R2,x01R3 | 02.12.04.00,02.12.02.00, 0212000B0AAAAAA, 0212000B0AAABAB, 0212000B0AAACAC, 0212000C0AAAAAA, 0212000C0AAABAB, 0212000C0AAACAC, 0212000M0AAAAAA, 0212000M0AAABAB, 0212000X0AAAAAA, 0212000X0AAABAB, 0212000X0AAADAD, 0212000Y0AAAAAA, 0212000Y0AAABAB, 0212000Y0AAADAD, 02120200, 02.12.02.00.00, 02120400, 02.12.04.00.00 | 134489001, 319996000, 319997009, 320000009, 320006003, 320012008, 320013003, 320014009, 320022002, 320023007, 320025000, 320029006, 320030001, 320031002, 320035006, 320036007, 320037003, 320041004, 408024009, 408036003, 408037007, 409108001, 414177002, 414178007, 414179004, 4580311000001109, 19722411000001106, 19722511000001105, 20528511000001106, 20528611000001105, 240705001000027108, 240715001000027105, 299275001000027104 |

Variable definitions constructed using ICD-9, ICD-10, OPCS-4, READ2 and CTV3 codes as well as self-report data fields with disease- or procedure-specific codes between brackets are shown. *: We additionally removed participants with glycated haemoglobin (HbA1c) ≥ 48 mmol/mol at baseline were removed from the Cox regression analysis. Abbreviations: CTV3, Clinical Terms Version 3; ICD, International Classification of Diseases; OPCS, Office of Population, Censuses and Surveys: Classification of interventions and Procedure.

**Table S2. Censoring dates for participants by region**

| Region | Censoring date |
|---|---|
| England | 30th September 2021 |
| Scotland | 31st July 2021 |
| Wales | 28th February 2018 |

**Table S3. Multivariable Cox proportional hazard analysis of Lp(a) concentration on type 2 diabetes**

| Model | Covariates | HR | 95% CI | *p* | Model | HR | 95% CI | *p* |
|---|---|---|---|---|---|---|---|---|
| ***all participants*** | | | | | | | | |
| | very low Lp(a) | | | | very high Lp(a) | | | |
| Model 1 | Age, Sex | 1.17 | 1.12 - 1.23 | <0.001 | Model 1 | 1.07 | 1.00 - 1.13 | 0.028 |
| Model 2 | Model 1 + BMI and hypertension | 1.11 | 1.06 - 1.17 | <0.001 | Model 2 | 1.00 | 0.95 - 1.06 | 0.867 |
| Model 3 | Model 2 + LDL-C + HDL-C | 1.10 | 1.04 - 1.15 | <0.001 | Model 3 | 1.06 | 1.00 - 1.12 | 0.072 |
| Model 4 | Model 3 + triglycerides | 1.06 | 1.01 - 1.12 | 0.016 | Model 4 | 1.08 | 1.02 - 1.15 | 0.009 |
| ***participants not on statin*** | | | | | | | | |
| | very low Lp(a) | | | | very high Lp(a) | | | |
| Model 1 | Age, Sex | 1.16 | 1.10 - 1.23 | <0.001 | Model 1 | 0.91 | 0.84 - 0.98 | 0.019 |
| Model 2 | Model 1 + BMI and hypertension | 1.10 | 1.04 - 1.16 | <0.001 | Model 2 | 0.90 | 0.83 - 0.98 | 0.015 |
| Model 3 | Model 2 + LDL-C + HDL-C | 1.10 | 1.03 - 1.16 | 0.003 | Model 3 | 0.94 | 0.86 - 1.02 | 0.125 |
| Model 4 | Model 3 + triglycerides | 1.06 | 1.00 - 1.13 | 0.055 | Model 4 | 0.97 | 0.89 - 1.05 | 0.442 |
| ***participants on statin*** | | | | | | | | |
| | very low Lp(a) | | | | very high Lp(a) | | | |
| Model 1 | Age, Sex | 1.25 | 1.15 - 1.35 | <0.001 | Model 1 | 0.94 | 0.86 - 1.02 | 0.112 |
| Model 2 | Model 1 + BMI and hypertension | 1.18 | 1.08 - 1.28 | <0.001 | Model 2 | 0.95 | 0.88 - 1.03 | 0.253 |
| Model 3 | Model 2 + LDL-C + HDL-C | 1.18 | 1.08 - 1.28 | <0.001 | Model 3 | 0.99 | 0.91 - 1.08 | 0.771 |
| Model 4 | Model 3 + triglycerides | 1.14 | 1.05 - 1.24 | 0.003 | Model 4 | 1.02 | 0.93 - 1.11 | 0.711 |

Very low Lp(a): Lp(a) <3.8 nmol/L; very high Lp(a): Lp(a) >189 nmol/L; Reference: Lp(a)3.8-189 nmol/L. BMI: body mass index, CI: confidence interval, HDL-C: high density lipoprotein cholesterol, HR: hazard ratio, LDL-C: low-density lipoprotein cholesterol.

**Table S4. Multivariate logistic regression of Lp(a) concentration on non-alcoholic liver disease**

| Model | Covariates | HR | 95% CI | *p* | Model | HR | 95% CI | *p* |
|---|---|---|---|---|---|---|---|---|
| ***all participants*** | | | | | | | | |
| | very low Lp(a) | | | | very high Lp(a) | | | |
| Model 1 | Age, Sex | 1.53 | 1.43 - 1.64 | <0.001 | Model 1 | 0.89 | 0.80 - 0.98 | 0.023 |
| Model 2 | Model 1 + BMI and hypertension | 1.45 | 1.35 - 1.55 | <0.001 | Model 2 | 0.84 | 0.76 - 0.93 | 0.001 |
| Model 3 | Model 2 + LDL-C + HDL-C | 1.39 | 1.30 - 1.50 | <0.001 | Model 3 | 0.89 | 0.80 - 0.99 | 0.031 |
| Model 4 | Model 3 + triglycerides | 1.35 | 1.26 - 1.45 | <0.001 | Model 4 | 0.91 | 0.82 - 1.01 | 0.084 |
| ***participants not on statin*** | | | | | | | | |
| | very low Lp(a) | | | | very high Lp(a) | | | |
| Model 1 | Age, Sex | 1.58 | 1.46 - 1.72 | <0.001 | Model 1 | 0.78 | 0.68 - 0.90 | <0.001 |
| Model 2 | Model 1 + BMI and hypertension | 1.51 | 1.39 - 1.64 | <0.001 | Model 2 | 0.78 | 0.68 - 0.90 | <0.001 |
| Model 3 | Model 2 + LDL-C + HDL-C | 1.44 | 1.32 - 1.57 | <0.001 | Model 3 | 0.85 | 0.73 - 0.98 | 0.029 |
| Model 4 | Model 3 + triglycerides | 1.40 | 1.28 - 1.53 | <0.001 | Model 4 | 0.88 | 0.76 - 1.02 | 0.083 |
| ***participants on statin*** | | | | | | | | |
| | very low Lp(a) | | | | very high Lp(a) | | | |
| Model 1 | Age, Sex | 1.42 | 1.26 - 1.61 | <0.001 | Model 1 | 0.80 | 0.69 - 0.92 | 0.002 |
| Model 2 | Model 1 + BMI and hypertension | 1.34 | 1.18 - 1.51 | <0.001 | Model 2 | 0.83 | 0.71 - 0.96 | 0.012 |
| Model 3 | Model 2 + LDL-C + HDL-C | 1.31 | 1.15 - 1.50 | <0.001 | Model 3 | 0.85 | 0.73 - 1.00 | 0.047 |
| Model 4 | Model 3 + triglycerides | 1.27 | 1.12 - 1.45 | <0.001 | Model 4 | 0.87 | 0.75 - 1.02 | 0.084 |

Very low Lp(a): Lp(a) <3.8 nmol/L; very high Lp(a): Lp(a) >189 nmol/L; Reference: Lp(a)3.8-189 nmol/L. BMI: body mass index, CI: confidence interval, HDL-C: high density lipoprotein cholesterol, HR: hazard ratio, LDL-C: low-density lipoprotein cholesterol.

**Table S5. Association of very low Lp(a) concentration with diabetes stratified by self-reported ethnic groups**

| Ethnic group | Model | Covariates | HR | 95% CI | *p* | N | $N_{event}$ |
|---|---|---|---|---|---|---|---|
| White | | | | | | | |
| | Model 1 | Age, Sex | 1.24 | 1.17-1.31 | <0.001 | 318,960 | 9,493 |
| | Model 2 | Model 1 + BMI and hypertension | 1.17 | 1.10-1.24 | <0.001 | 317,477 | 9,420 |
| | Model 3 | Model 2 + LDL-C + HDL-C | 1.17 | 1.10-1.24 | <0.001 | 289,203 | 8,591 |
| | Model 4 | Model 3 + triglycerides | 1.13 | 1.07-1.21 | <0.001 | 289,169 | 8,589 |
| Asian | | | | | | | |
| | Model 1 | Age, Sex | 1.08 | 0.75-1.56 | 0.671 | 6,634 | 554 |
| | Model 2 | Model 1 + BMI and hypertension | 0.97 | 0.67-1.41 | 0.882 | 6,579 | 542 |
| | Model 3 | Model 2 + LDL-C + HDL-C | 1.06 | 0.71-1.57 | 0.791 | 5,982 | 489 |
| | Model 4 | Model 3 + triglycerides | 1.04 | 0.70 - 1.55 | 0.828 | 5,982 | 489 |
| Black | | | | | | | |
| | Model 1 | Age, Sex | 1.14 | 0.51-2.55 | 0.756 | 4,844 | 402 |
| | Model 2 | Model 1 + BMI and hypertension | 1.26 | 0.56-2.83 | 0.573 | 4,775 | 395 |
| | Model 3 | Model 2 + LDL-C + HDL-C | 0.68 | 0.22-2.12 | 0.507 | 4,388 | 364 |
| | Model 4 | Model 3 + triglycerides | 0.51 | 0.16-1.62 | 0.254 | 4,387 | 364 |
| Mixed | | | | | | | |
| | Model 1 | Age, Sex | 1.49 | 0.72-3.09 | 0.283 | 2,097 | 85 |
| | Model 2 | Model 1 + BMI and hypertension | 1.59 | 0.77-3.31 | 0.211 | 2,084 | 85 |
| | Model 3 | Model 2 + LDL-C + HDL-C | 1.31 | 0.57-3.03 | 0.524 | 1,893 | 76 |
| | Model 4 | Model 3 + triglycerides | N/A | | | | |
| Other | | | | | | | |
| | Model 1 | Age, Sex | 0.98 | 0.62-1.55 | 0.930 | 4,362 | 254 |
| | Model 2 | Model 1 + BMI and hypertension | 1.01 | 0.64-1.60 | 0.967 | 4,288 | 250 |
| | Model 3 | Model 2 + LDL-C + HDL-C | 0.93 | 0.56-1.52 | 0.760 | 3,913 | 231 |
| | Model 4 | Model 3 + triglycerides | 0.85 | 0.52-1.41 | 0.538 | 3,913 | 231 |

Very low Lp(a): Lp(a) <3.8 nmol/L; Reference: Lp(a)3.8-189 nmol/L. Models with <10 event-per-variable were not reported. BMI: body mass index, CI: confidence interval, HDL-C: high density lipoprotein cholesterol, HR: hazard ratio, LDL-C: low-density lipoprotein cholesterol, N: Number of participants in group, $N_{event}$: number of events for the group.

**Table S6. Association of very low Lp(a) concentration with non-alcoholic liver disease stratified by self-reported ethnic groups**

| Ethnic group | Model | Covariates | HR | 95% CI | *p* | N | $N_{event}$ |
|---|---|---|---|---|---|---|---|
| White | | | | | | | |
| | Model 1 | Age,Sex | 1.59 | 1.46-1.73 | <0.001 | 325,393 | 3963 |
| | Model 2 | Model 1 + BMI and hypertension | 1.50 | 1.38-1.63 | <0.001 | 323,847 | 3936 |
| | Model 3 | Model 2 + LDL-C + HDL-C | 1.45 | 1.33-1.58 | <0.001 | 295,025 | 3595 |
| | Model 4 | Model 3 + triglycerides | 1.41 | 1.29-1.54 | <0.001 | 294,988 | 3594 |
| Asian | | | | | | | |
| | Model 1 | Age,Sex | 1.56 | 0.79-3.08 | 0.204 | 7239 | 110 |
| | Model 2 | Model 1 + BMI and hypertension | 1.47 | 0.74-2.91 | 0.273 | 7168 | 107 |
| | Model 3 | Model 2 + LDL-C + HDL-C | 1.07 | 0.47 - 2.46 | 0.870 | 6524 | 98 |
| | Model 4 | Model 3 + triglycerides | 1.03 | 0.45 - 2.37 | 0.943 | 6524 | 98 |
| Black | | | | | | | |
| | Model 1 | Age,Sex | 2.16 | 0.53-8.84 | 0.283 | 5242 | 66 |
| | Model 2 | Model 1 + BMI and hypertension | 2.39 | 0.58-9.82 | 0.226 | 5166 | 65 |
| | Model 3 | Model 2 + LDL-C + HDL-C | N/A | | | | |
| | Model 4 | Model 3 + triglycerides | N/A | | | | |
| Mixed | | | | | | | |
| | Model 1 | Age,Sex | N/A | | | | |
| | Model 2 | Model 1 + BMI and hypertension | N/A | | | | |
| | Model 3 | Model 2 + LDL-C + HDL-C | N/A | | | | |
| | Model 4 | Model 3 + triglycerides | N/A | | | | |
| Other | | | | | | | |
| | Model 1 | Age,Sex | 1.90 | 1.03- 3.5 | 0.040 | 4608 | 85 |
| | Model 2 | Model 1 + BMI and hypertension | 1.91 | 1.03-3.52 | 0.039 | 4523 | 84 |
| | Model 3 | Model 2 + LDL-C + HDL-C | 1.54 | 0.79-3.00 | 0.204 | 4135 | 79 |
| | Model 4 | Model 3 + triglycerides | N/A | | | | |

Very low Lp(a): Lp(a) <3.8 nmol/L; Reference: Lp(a)3.8-189 nmol/L. Models with <10 event-per-variable were not reported. BMI: body mass index, CI: confidence interval, HDL-C: high density lipoprotein cholesterol, HR: hazard ratio, LDL-C: low-density lipoprotein cholesterol, N: Number of participants in group, $N_{event}$: number of events for the group.

#### Figure S1. Mendelian Randomization methods and sensitivity analyses

Assumption IV1 (the variant is predictive of the exposure) (**A**) can be assessed using the F-statistics when using a two-sample univariable MR-inverse-variance weighted (IVW) analysis. In case the MR-Egger approach is used, weak instrument bias can be assessed using the $I^2_{GX}$ index. Assumption IV2 (**B**; the variant is independent of any confounding factors of the exposure—outcome association) cannot be proven. However, several analyses were undertaken to disprove it or estimate a causal effect in the presence of violation of the assumption. **B1**) Confounding. Confounding is, in theory, limited by the second law of Mendel, which states that genetic variants for different traits are inherited independently. To further limit confounding, the exposure GWAS was corrected using standard methodology and the samples of the exposure and outcome GWAS did not have any overlap. **B2**) Horizontal pleiotropy. MR-PRESSO and leave-one out analyses exclude some, but not as many as median and mode-based analyses, SNPs from the MR estimate using a data-driven or systematic approach. They are especially useful to see whether the results are robust to a few horizontally pleiotropic outliers. Assumption IV3 (**C**; the variant is conditionally independent of the outcome given the exposure and the confounding factors) can also not be proven. We undertook the following steps to detect its violation or to estimate a causal effect in the presence of its violation. First, (**C1**) was explored using the Rücker framework which assesses pleiotropy distribution using heterogeneity in MR-IVW (Cochran Q), MR-Egger (Rücker Q') and by calculating their difference (Q-Q'). The results can be used to move between standard MR methods (MR-IVW fixed and random effects, MR-Egger). Additionally, the $I^2$ index is calculated to assess potential heterogeneity. **C2**) was assessed using Steiger filtering, which removes variants from the analysis if they are more strongly associated with the outcome than with the exposure. Optionally, it is possible to perform a bi-directional MR to evaluate whether the exposure causes the outcome, or the outcome causes the exposure. This was not performed in the present study.

Finally, weighted median and mode-based analyses were performed as sensitivity analyses which are more robust in terms of violation of assumptions IV2 and IV3. Since these methods allow for half or the majority of the SNPs to be invalid, they have a natural robustness to variants with outlying ratio estimates, and so are not as affected by the presence of a small number of pleiotropic variants as the IVW and MR-Egger methods.

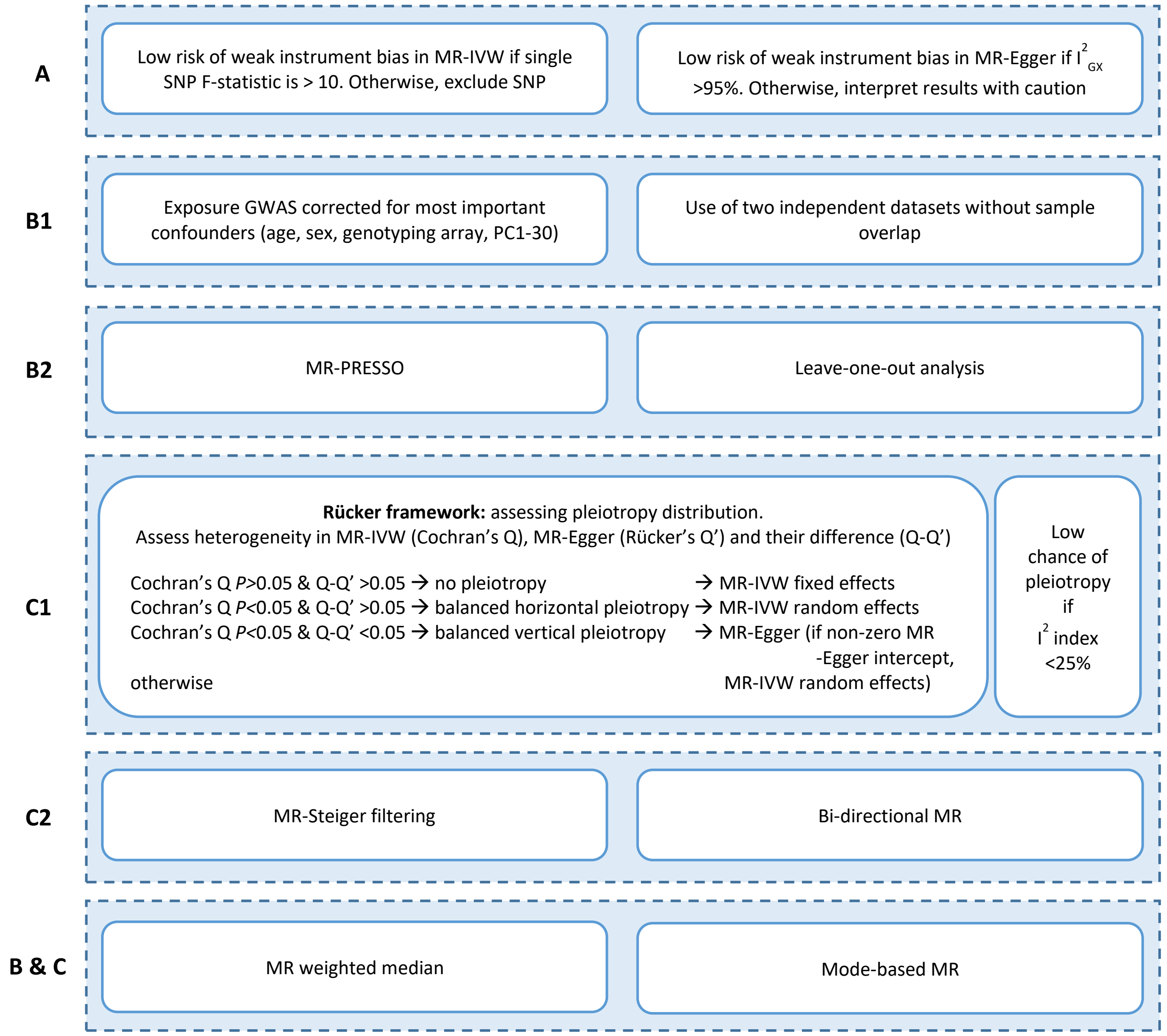

**Figure S2. Forest plot illustrating the unadjusted and adjusted hazard ratios of very high Lp(a) values at baseline**

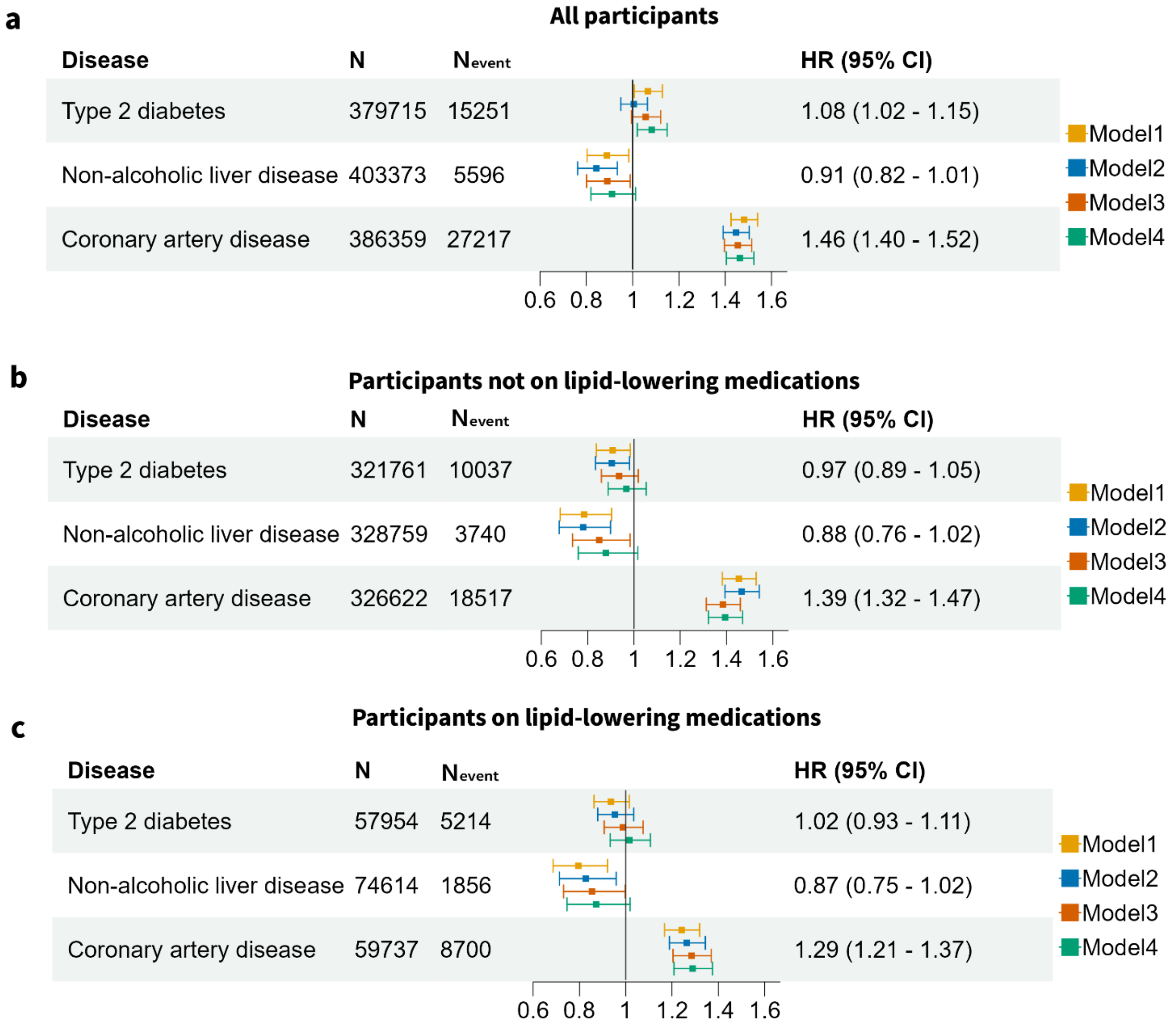


Hazard ratios for diseases in **(a)** all participants, **(b)** participants not using lipid-lowering medications and **(c)** participants using lipid-lowering medications in model 4. Model 1: adjusted for age and sex; Model 2: Model 1 + body mass index and hypertension; Model 3: Model 2 + low- density lipoprotein cholesterol and high-density lipoprotein cholesterol; Model 4: Model 3 + triglycerides. Hazard ratios (HR) with 95% confidence intervals (CI) of Model 4 are shown for new-onset T2D, NAFLD and CAD. N: Number of participants, $N_{event:}$ number of events.

**Figure S3. Forest plot illustrating the unadjusted and adjusted hazard ratios of inverse rank normalized Lp(a) values at baseline**

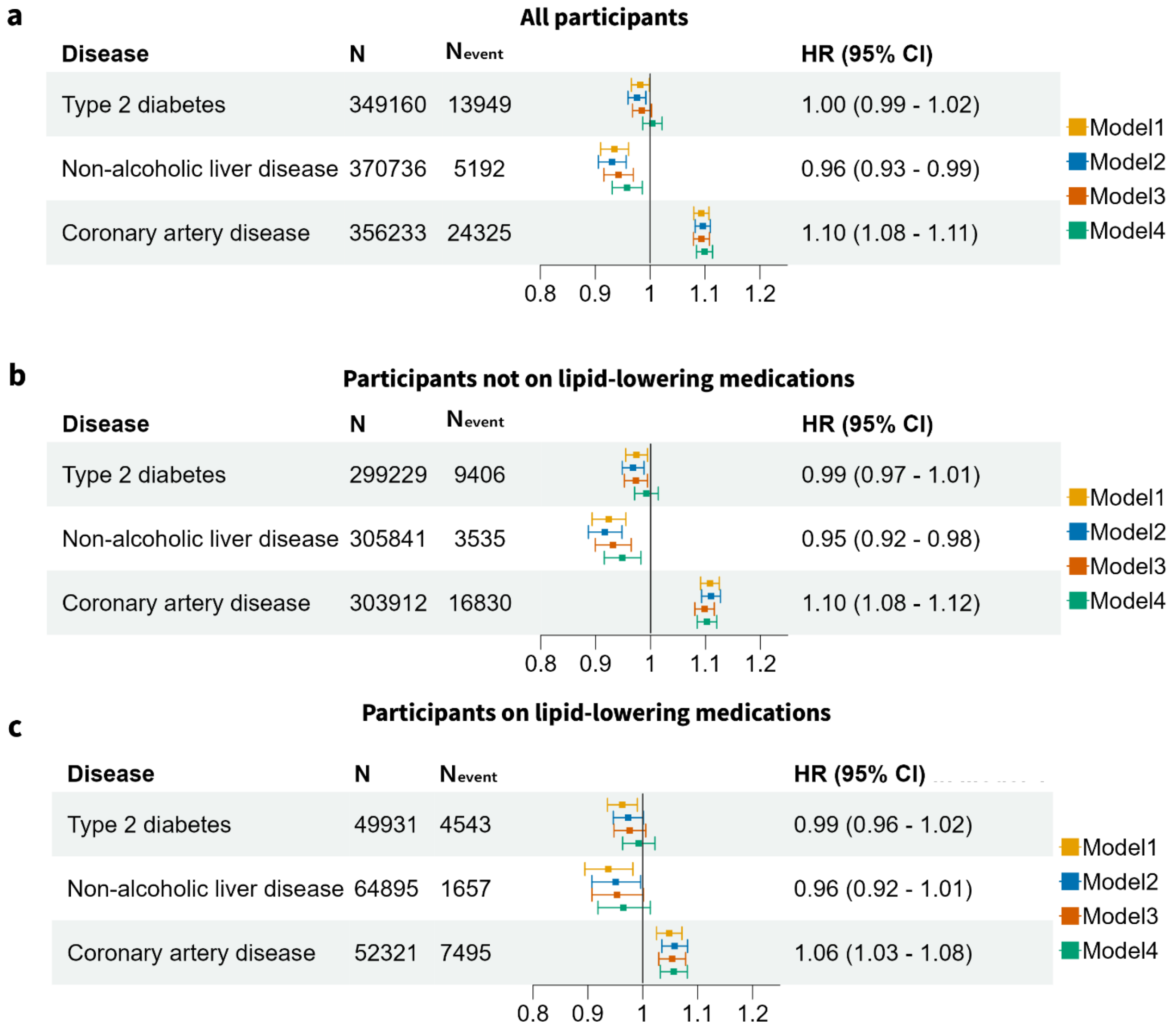


Hazard ratios for diseases in **(a)** all participants, **(b)** participants not using lipid-lowering medications and **(c)** participants using lipid-lowering medications in model 4. Model 1: adjusted for age and sex; Model 2: Model 1 + body mass index and hypertension; Model 3: Model 2 + low- density lipoprotein cholesterol and high-density lipoprotein cholesterol; Model 4: Model 3 + triglycerides. Hazard ratios (HR) with 95% confidence intervals (CI) of Model 4 are shown for new-onset T2D, NAFLD and CAD. N: Number of participants, $N_{event}$: number of events.
